# The impact of headache disorders on COVID-19 survival: a world population-based analysis

**DOI:** 10.1101/2021.03.10.21253280

**Authors:** Robert E. Shapiro, Víctor J. Gallardo, Edoardo Caronna, Patricia Pozo-Rosich

**Author notes:** Correspondence to: Patricia Pozo-Rosich, Ps. Vall d’Hebron 119-129, 08035 Barcelona – Spain. These authors contributed equally to this work (Robert E. Shapiro, Víctor J. Gallardo).

## Abstract

**Importance:** COVID-19 has not impacted people or countries uniformly. This disparity has prompted investigations to identify clinical and genetic predictors of COVID-19 mortality. Headache, a COVID-19 symptom, has been associated with positive disease prognosis. It is logical to consider whether primary headache disorders, among the most prevalent and disabling diseases globally, may also be associated with reduced viral mortality and thereby may have arisen as adaptive host defences

**Objective:** To study the relationship between COVID-19 mortality and primary headache disorders.

**Main outcome measure:** Using a generalized additive model regression (GAM), we analysed data across 171 nations to identify variables which impact COVID-19 mortality rates (demographics, national wealth and government effectiveness, pandemic management indexes, latitude of the country’s capital, prevalence of headache disorders and other diseases). We performed similar analyses of seasonal influenza mortality. Separately, we meta-analysed studies of COVID-19 inpatient survival reporting headache, using PRISMA guidelines.

**Results:** In the global population-level analysis, we observed that a higher prevalence of headache disorders was associated with a higher COVID-19 mortality rate, and represented the main variable contributing to differences in COVID-19 mortality rates between countries (37.8%; *F* value=10.68). By contrast, we observed a negative trend between the prevalence of headache disorders and influenza death rates. Controversially, when considering headache as a symptom of COVID-19, in the 48 meta-analysed studies we observed a significantly higher risk ratio of survival (RR:2.178 [1.882-2.520], p<0.0001) among COVID-19 inpatients with headache.

**Conclusions and Relevance:** Headache as a primary disorder is more prevalent in nations with higher COVID-19 mortality, whereas headache as a COVID-19 symptom is associated with enhanced survival. Further studies should clarify whether primary headache disorders represent a risk factor for mortality for COVID-19 or, rather, whether this association reflects evolutionary adaptive processes to enhance survival that, in the case of COVID-19, are insufficiently protective.

## Introduction

The COVID-19 pandemic has not impacted global populations uniformly. Some regions have reported significantly lower COVID-19 prevalence and death rates compared to other regions (i.e. tropical Africa and East Asia vs Europe and North America).^1^ The bases for these disparities are unclear, but may include social factors, reporting methods, demographic, environmental, geographic, intrinsic health, or genetic determinants. Some traits found to be associated with increased COVID-19 mortality include advanced age, male sex, deprivation, and chronic diseases.^2^ Moreover, several human genomic variants have been linked to variations in COVID-19 outcomes.^3-5^ On the other hand, COVID-19 traits associated with a *positive* disease prognosis have received less attention from investigators. One such trait may be headache.

We studied a prospective, consecutive cohort of 130 patients admitted for COVD-19 to a Spanish hospital, and found that inpatients reporting headache on admission had COVID-19 symptoms for one less week than inpatients not reporting headache.^6^

We sought to confirm the observation that headache, as a COVID-19 symptom, is a putative marker of favourable COVID-19 clinical outcomes, specifically mortality. We now report a meta-analysis of 48 inpatient studies which included the presence or absence of headache as a COVID-19 symptom and reported COVID-19 mortality rates. Separately, we further investigated the possible relationship between COVID-19 death rates and the prevalence of primary headache disorders across 171 national populations.

## Methods

### COVID-19 mortality: headache as a COVID-19 symptom

#### Meta-analysis

##### Search strategy, study selection and data extraction

We conducted a systematic literature search of PubMed (April 1, 2020 to December 22, 2020) to identify all COVID-19 clinical inpatient series in accordance with the PRISMA guideline.^7^ We also included 6 studies published between December 2019 and March 2020 from a previous meta-analysis.^8^ We excluded review articles, opinion articles, case-reports, preprint server articles, and studies performed either on populations <18 years old or animal models (See Supplementary Methods and Supplementary Table 1).

##### Data analysis

Random-effects pooling models were computed in order to estimate the effect size of the following binary outcome data: presence of headache in survived vs. non-survived COVID-19 cohorts.^9^ Pooled headache prevalence and 95% confidence intervals (CIs) were presented from selected publications. Risk ratio (RR) with 95% CIs were used to estimate the risk of experiencing headache in both COVID-19 cohorts: survivors and non-survivors.^10^ RR was computed using the Mantel-Haenszel method.^11^ Headache prevalence and RR from each publication were reported using forest plots. Headache RR was also analysed in different COVID-19 subgroups. Between-study heterogeneity was assessed using the I^2^ statistic and the Cochran’s Q-test for statistical significance.^12^ Outlier publications were discarded in the sensitivity analysis in order to check the robustness of our results. We repeated the same analysis for the other COVID-19 symptoms and patients’ comorbidities collected, although not all publications recorded the same COVID-19 symptomatology or patients’ comorbidities. Hence, we analysed their pooled prevalence and risk ratio in publications where headache was reported.

### COVID-19 mortality-related factors: headache disorders

#### Data collection

We downloaded the cumulative COVID-19 deaths per million population for each country from the start of the pandemic until the start of the global COVID-19 vaccination campaign (December 14, 2020). Then, for each country, we downloaded variables describing their population and demographics, national wealth and government effectiveness, pandemic management indexes, latitude of the country’s capital, and prevalence of headache disorders and other diseases. Source data for each variable are described in the Supplementary Table 2. Only countries with data available both for the prevalence of headache disorders and other diseases, and for COVID-19 death rates were included.

#### Data modelling and analysis

We consider the cumulative COVID-19 deaths per million population as the response variable and we evaluated other possible influencing factors as explanatory variables in a generalized additive model (GAM). In the single-factor model, we evaluated all explanatory variables individually and we measured their significance. All statistically significant variables from the univariate analysis were then considered in the multi-factor model in order to assess their independence for the response variable. Spearman correlation was computed in order to quantify the degree of correlation between each predictor. Predictors with correlation coefficient greater than 70% were not included simultaneously in the multi-factor model. Hence, we used the forward selection in R to derive the final GAM for the analysis.

More details are included in the Supplementary Methods.

## Results

### COVID-19 mortality: headache as a COVID-19 inpatient symptom

The meta-analysis included a total of 48 full-text peer-reviewed publications of COVID-19 inpatient mortality studies, that also reported headache as a COVID-19 symptom (Supplementary Fig. 1 and Supplementary Table 3).

Although there was statistically significant heterogeneity between studies, the overall pooled prevalence of headache as a symptom among COVID-19 inpatients was 9.7% [7.8%-12-0%] (Fig. 1A, Supplementary Table 4).

**Figure 1.**
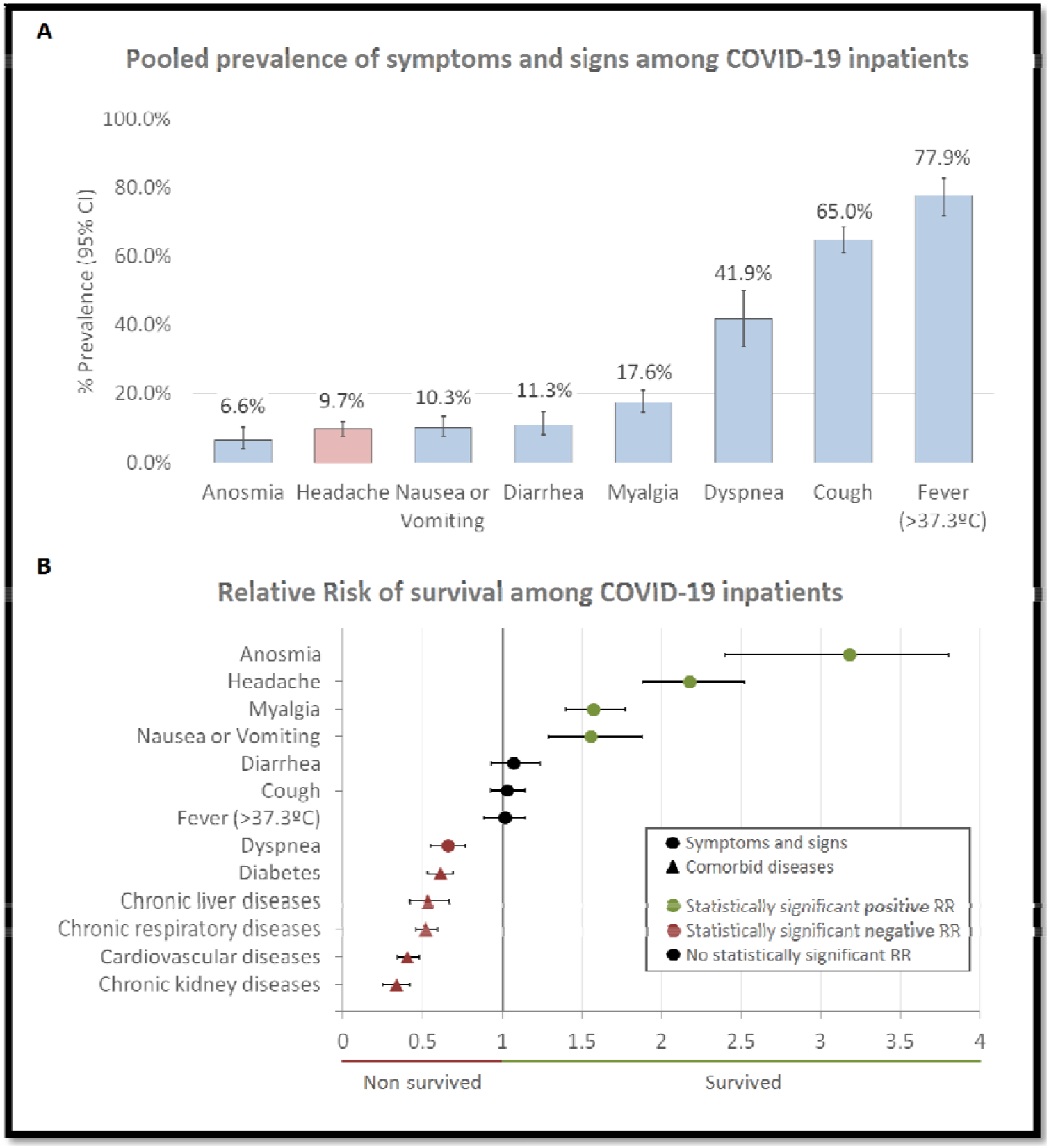
Pooled prevalence of symptoms and signs among COVID-19 inpatients (A) and relative risk of survival for COVID-19 inpatients relative to their symptoms, signs and comorbid diseases (B). Abbreviations: CI: Confidence Interval; RR: Relative risk factors.

Regarding the risk of headache relative to mortality, we observed a higher risk ratio of headache among COVID-19 inpatients who survived, compared to those who did not (RR: 2.178 [1.882-2.520], p<0.0001), excluding outliers and over-influencer studies (I^2^ = 16%, p=0.175) (Fig. 2). Notably, some other COVID-19 symptoms were also associated with significantly higher RR for survival, including anosmia (RR: 3.183 [2.397-4.226]), myalgia (RR: 1.574 [1.398-1.773]), and nausea or vomiting (RR: 1.558 [1.292-1.879]), whereas all comorbid diseases studied were associated with COVID-19 non-survival (RR < 1) (Fig. 1B, Supplementary Tables 5 and 6). Finally, we performed sensitivity analyses of headache RR, and consistently observed higher RR of headache among COVID-19 inpatients who survived (Supplementary Table 7). Moreover, risk of headache did not exhibit statistically significant publication bias following visual inspection and Egger’s test (Supplementary Fig. 2).

**Figure 2.**
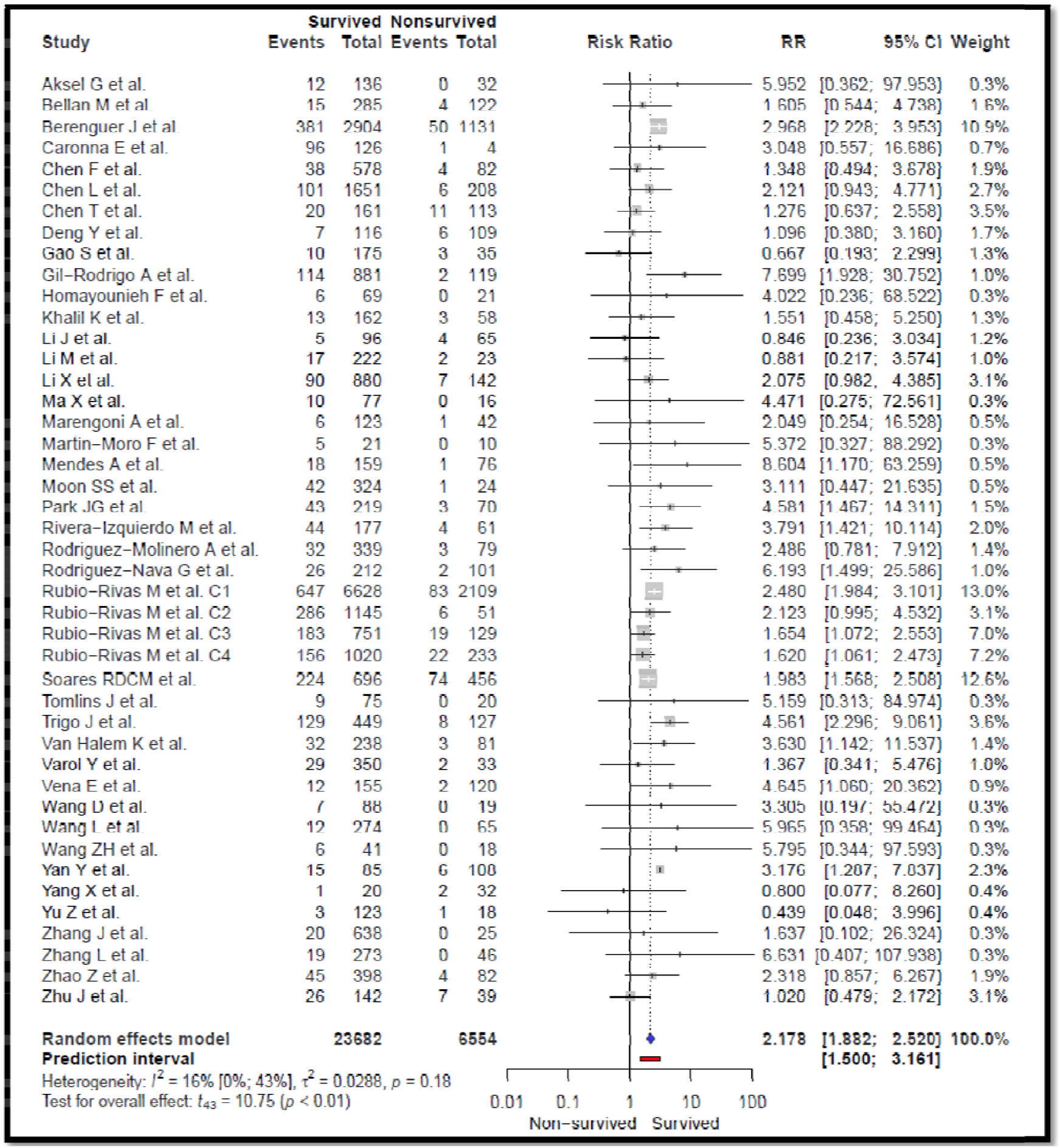
Risks of headache among survived (recovered or discharged) vs non-survived COVID-19 inpatients. From the 48 selected studies, 7 were removed in order to obtain a homogeneous effect between studies. For citations see Supplementary Table 3 and Supplement meta-analysis references.

### COVID-19 mortality-related factors: headache disorders

For the 171 countries for which data were available, the COVID-19 death rate was not uniform amongst all world regions (Fig. 3G-3H); Europe, North America and South America were the most affected continents. We observed a statistically significant trend towards higher COVID-19 mortality rate in countries with higher prevalence of headache disorders (Fig. 3A), or chronic respiratory diseases (Fig. 3E). Conversely, countries with higher prevalence of cirrhosis and chronic liver diseases, were associated with lower COVID-19 mortality rates (Fig. 3D). We found significant correlations between prevalence of different primary headache disorders and higher COVID-19 mortality, as well as with higher latitude or with higher population median age (Fig. 4 and Supplementary Fig. 3).

**Figure 3.**
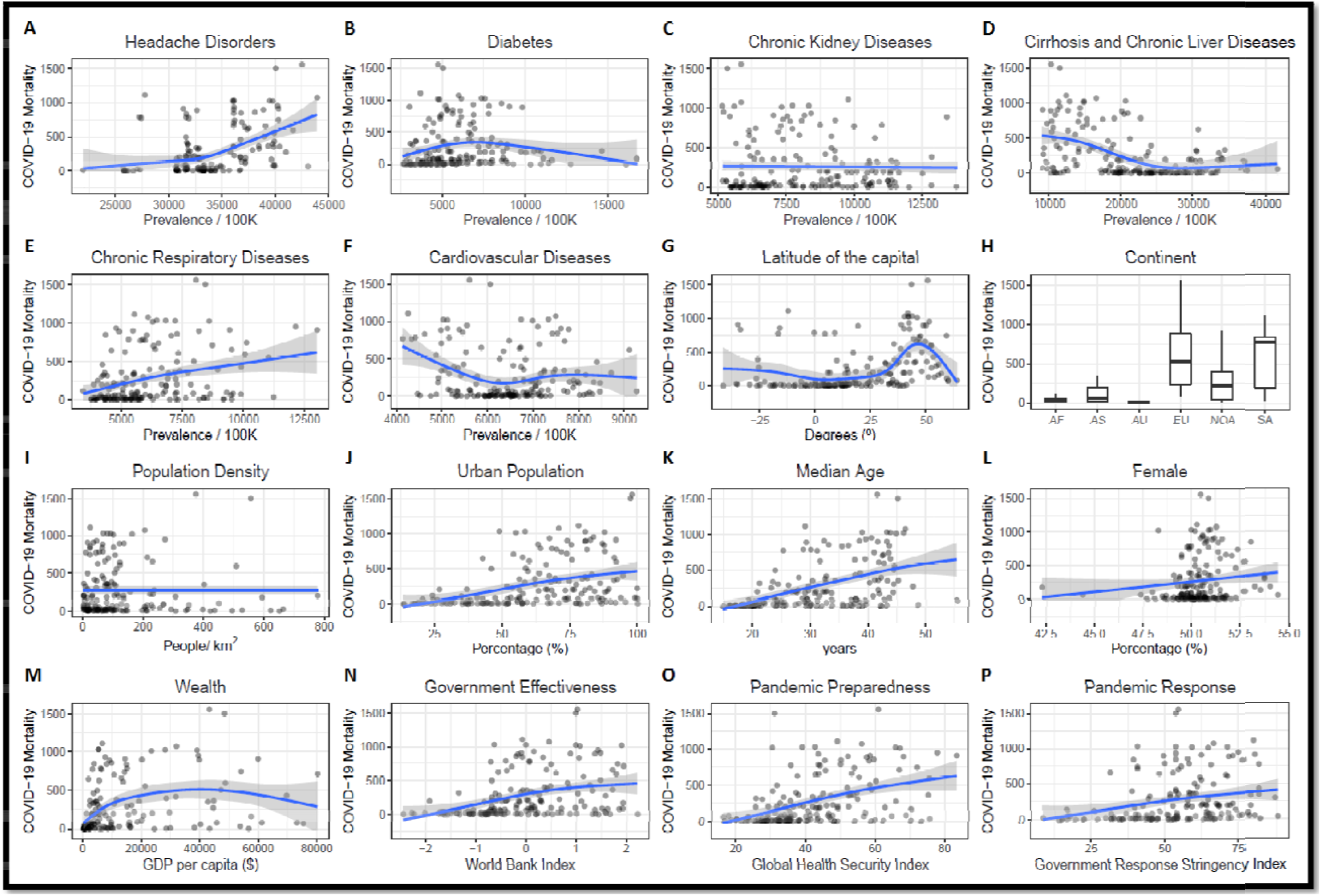
COVID-19 mortality (per million population) distribution across 171 nations based on geographical distribution and diseases prevalence (according to 2019): (A) Headache disorders; (B) Diabetes mellitus; (C) Chronic kidney diseases; (D) Cirrhosis and Chronic liver diseases; (E) Chronic respiratory diseases an Cardiovascular diseases; (G) Capital’s latitude; (H) Continents; (I) Population density; (J) Urban population rate; (K) Population’s median age; (L) Population’s fe rate; (M) National wealth; (N) Government effectiveness; (O) Pandemic preparedness; (P) Pandemic response Abbreviations: GBD: Global Burden of Diseases; AF: Africa; AS: Asia; AU: Australia; EU: Europe; NOA: North-America; SA: South-America.

**Figure 4.**
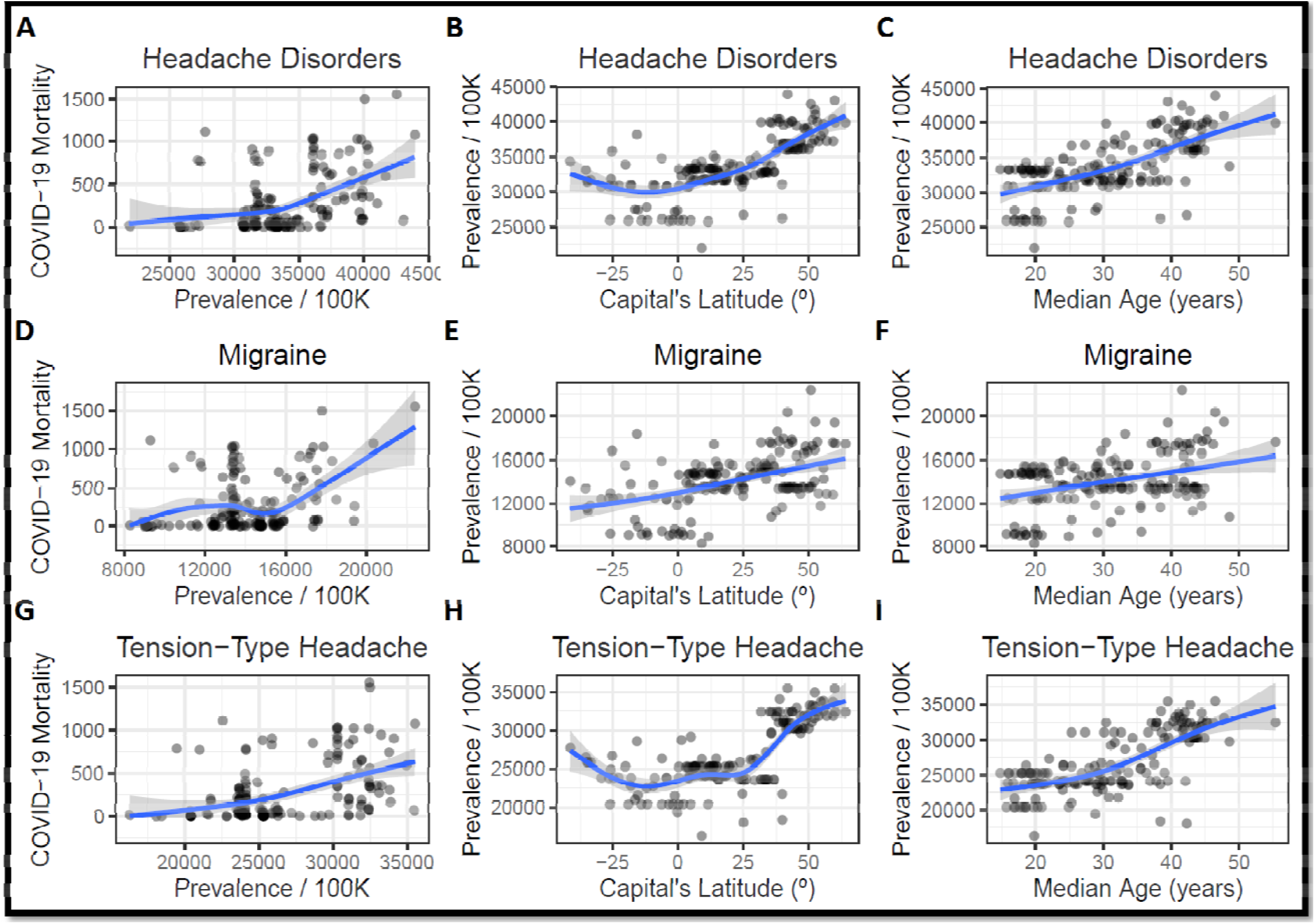
COVID-19 mortality (per million population) distribution across 171 nations based on headache disorders prevalence (according to GBD2019) latitudinal distribution, and country’s median age: (ABC) All headache disorders prevalence considered in the GBD2019; (DEF) Migraine prevalence and (GHI); Tension-type headache prevalence. GBD: Global Burden of Disease

To study the effect of each variable on COVID-19 mortality, we conducted a GAM regression. Supplementary Table 8 shows the statistical significance and the deviance explained for each predictor, individually, on COVID-19 mortality. The deviance explained represents the percentage of the total variance explained or determined by the available predictors and is often used to evaluate the fit of the model. In the single-factor model, the highest deviance explained was observed for headache disorders prevalence (including migraine and tension-type headache) which contributed 37.8% (with an *F* value of 10.68) of the differences in COVID-19 mortality between countries. However, migraine prevalence alone contributed 32.0% (*F* value = 8.38) (Fig. 5, Supplementary Table 8). This indicates that prevalence of headache disorders, and specifically migraine, are the most significant factors, yet identified, that may drive differences in COVID-19 mortality between countries.

**Figure 5.**
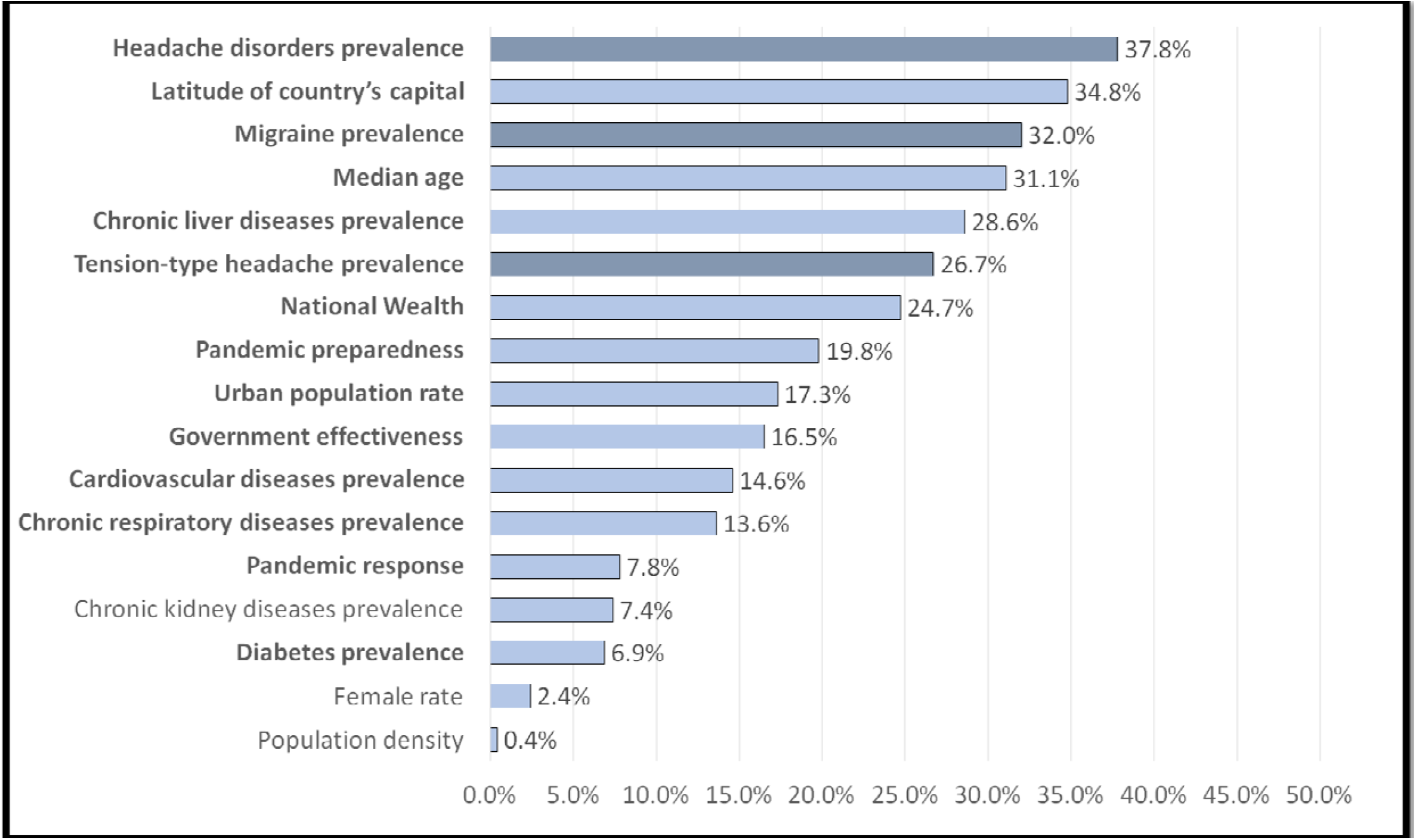
Deviance explained (%) by the variables associated with COVID-19 mortality across 171 nations in the generalized additive single-factor model The deviance explained represents the percentage of the total variance determined by the predictors, individually (single-factor model), and it is often used to evaluate the fit of the model. A larger value indicates that the model fits better. In the Supplementary Table 7 is shown all model’s coefficients and statistical significance. In **bold** are statistically significant (P < 0.05) smoothed variables.

Regarding other potential predictors of COVID-19 mortality rates, all statistically significant correlations with evaluated explanatory variables were positive, except the prevalence of chronic liver diseases, that is negative (Fig. 3D and Supplementary Fig. 3). Notably, and paradoxically, public health and social measures (PHSM) (i.e., national wealth, government effectiveness, pandemic preparedness, pandemic responses) were among the factors *positively* associated with higher mortality rates, suggesting limited impacts of these factors (Fig. 3M-3P, Supplementary Fig. 3). Based on the single factor model, statistically significant variables were used in a multi-factor GAM where migraine prevalence, urban population, national wealth, and latitude of the country’s capital were the four statistically significant factors with an adjusted R^2^ of 0.591 and 65.5% of deviance explained (Supplementary Table 9).

We also conducted a multi-factor GAM grouping countries by tropical vs non-tropical location to control for geographic latitude (Supplementary Table 10). 96.6% of COVID-19 mortality in tropical countries could be explained by seven variables (migraine, median age, national wealth, pandemic preparedness, urban population rate, chronic kidney diseases prevalence, and cardiovascular diseases prevalence), where wealth and pandemic preparedness were again, paradoxically, positive associations. Among non-tropical countries, only 68.3% of COVID-19 mortality was explained by all the indices analysed.

Finally, considering the significant association between COVID-19 mortality and prevalence of headache disorders, we tested if such association was present for seasonal influenza, a more common viral infection. By contrast to COVID-19, for the 171 selected countries, we observed a statistically significant negative correlation between 2019 influenza death rate and all the variables considered in the analysis, with the exception of chronic liver diseases that it was positive. Likewise, no statistically significant correlation was found between Influenza mortality and chronic kidney diseases prevalence and cardiovascular diseases prevalence (Supplementary Fig. 4). The prevalence of the comorbid diseases (including headache disorders, diabetes and chronic kidney and liver diseases) and latitude were independently associated with Influenza mortality in the generalized additive multi-factor model (Supplementary Table 11).

## Discussion

In light of our prior observations that the symptom of headache was associated with reduced length of COVID-19 disease^6^, we pursued two complementary analyses to assess whether headache may reflect a host viral defensive process.

First, we performed a meta-analysis of all 48 published COVID-19 inpatient mortality studies which captured headache as a symptom. This analysis indicates that inpatients that experience headache in the setting of the SARS-CoV-2 infection, are approximately twice as likely to survive, compared to those without headache.

Headache is a common concomitant symptom of systemic infections, particularly viral infections.^13^ COVID-19 is no exception, as recognized by the CDC at the outset of the pandemic.^14^ Headache, in the setting of COVID-19, may be a marker of host defensive responses to enhance survival. Multiple pathophysiological mechanisms might mediate these effects. The COVID-19 cytokine storm is associated with pulmonary inflammation and / or vasoconstriction mediated, in part, by interleukin 6 (IL-6).^15^ We had previously observed that IL-6 levels were lower and more stable in COVID-19 inpatients that reported headache.^6^ Circulating levels of calcitonin gene-related peptide (CGRP) levels are reportedly reduced during COVID-19^16^, though it is unclear whether this is pathological or compensatory. By contrast, CGRP is elevated during migraine attacks^17^, and headache during COVID-19 might reflect a host response that includes raising CGRP levels. Anti-CGRP medications are FDA-approved for migraine treatment^18^, and an anti-CGRP drug is under trial as a potential COVID-19 therapy^19^; it remains to be seen whether it may affect COVID-19 outcomes. The SARS-CoV-2 virus binds to the angiotensin converting enzyme 2 (ACE2) receptor on cell surfaces to gain cellular entry.^20^ The renin-angiotensin system is reported to be altered in people with migraine, and drugs that inhibit ACE1 or block angiotensin receptors are prescribed to prevent migraine attacks.^21^ Finally, vitamin D deficiency may be associated with COVID-19 mortality^22^, and vitamin D has been proposed as a migraine therapeutic.^23^

Collectively, these data suggest that headaches secondary to a SARS-CoV-2 infection may share certain pathophysiological mechanisms with primary headache disorders, including migraine. Therefore, if host responses that mediate headache as a COVID-19 symptom to enhance survival, it is logical to consider whether primary headache disorders may also be associated with reduced viral mortality.

Headache disorders are among the most prevalent and burdensome diseases.^24^ Migraine is highly prevalent (global mean 15%, range 7 – 23%), highly heritable, has onset in adolescence / early adult-hood, and affects three times more women than men.^25^ Taken together, it has been proposed that adaptive pressures may have selected for migraine susceptibility alleles to improve fitness. For example, the TRPM8 channel is the only known cold nociceptor, and specific TRPM8 alleles are associated both with increased cold sensitivity and heightened genetic risk of migraine expressed in a latitudinal gradient.^26^ Similarly, we hypothesize that the prevalence of headache disorders may also have increased in populations under specific viral selection pressures.

We therefore conducted an analysis of multiple indices across 171 national populations and observed, surprisingly, that higher population prevalence of headache disorders is strongly associated with *higher* COVID-19 death rates. The prevalence of headache disorders, particularly migraine, were principal factors explaining differences in distribution of elevated COVID-19 mortality across countries.

In interpreting these findings, we sought to ensure that headache disorders are not confounders of other factors involved in COVID-19 mortality. For example, we observed that COVID-19 population death rates also increase with distance from the equator. To mediate this effect, it has been proposed that climatic factors, particularly available sunlight and vitamin D, may influence either viral host defences or viral virulence, including that of SARS-CoV-2.^27^ As noted, our data and those of other studies^24^, show that the prevalence of headache disorders increases with higher latitude. However, in our analyses comparing tropical countries separately from non-tropical countries, migraine remains one of the main factors related to COVID-19 mortality (Supplementary Table 10).

Further, we sought to determine whether headache disorders are also associated with mortality due to more common and less virulent viral pathogens than SARS-CoV-2. We found that higher population prevalence of headache disorders is associated with reduced death rates due to influenza, by contrast to higher death rates due to SARS-CoV-2. This disparity suggests that headache disorders may indeed be a marker of enhanced host pathophysiological defences to viruses, but perhaps far less robustly to SARS-CoV-2 than to influenza viruses. Note, however, that a limitation of our analysis is that we could not obtain data to control for national variations in influenza vaccination rates.

The hypothesis that headache, as a *symptom*, may confer a competitive advantage in enhancing host defences from viral infections appears to be at odds with our finding that higher population prevalence of primary headache *disorders* is associated with increased COVID-19 mortality. This puzzle might be reconciled by consideration of the genetics of headache disorders. That is, susceptibility alleles for highly polygenic headache disorders (e.g. migraine) may have been selected as adaptive responses to coronaviruses, rather than that headache disorders directly increase the risk of COVID-19 death.^28^ In the case of novel SARS-CoV-2 infection, headache disorders might be insufficiently protective to markedly reduce COVID-19 death rates at a population level. In this context, neither host responses (i.e. headache disorders), nor social responses (i.e. PHSM), might be sufficient countermeasures to the high virulence of SARS-CoV-2.

Evidence is emerging that selection pressures that differ between Africa and Europe may have influenced the expression of COVID risk-altering human alleles. Zeberg and Pääbo have recently reported that the risk of developing clinically more severe COVID-19 is linked to chromosome 3 genetic variations, whereas the risk of developing clinically less severe COVID-19 is linked to chromosome 12 genetic variations.^5,29^ Moreover, these risk-altering haplotypes are of Neanderthal origin, and are more prevalent in European populations, compared to populations where COVID-19 death rates are relatively lower (i.e. sub-Saharan Africa, China). Similarly, early humans may have increased the frequency of existing viral defensive alleles, such as prevalent low-effect migraine susceptibility alleles, in response to selection pressures of severe coronavirus epidemics in northern climates. Genetic factors might thereby help explain why headache disorders, including migraine, emerged in our analyses as a main determinant of the differences in COVID-19 mortality across countries.

This study has significant strengths and limitations. The study combines and compares (1) analyses of meta-analytic symptoms and outcome data from 48 studies of COVID-19 inpatients, with (2) population level data from 171 nations regarding COVID-19 death rates. However, in doing so, we have relied on data from numerous sources, the accuracy of which we cannot independently confirm. Further, our observations of a latitudinal gradient effect for COVID-19 population mortality is strongly influenced by data from tropical African countries some of which are known to under-report death rates.^30^ Further, the higher number of tropical African countries (41), compared to tropical non-African countries (37), included in our latitudinal analyses also may introduce a bias towards effects of data from Africa. Mitigating these concerns are (1) our sub-analyses that show that a latitudinal gradient for COVID-19 death rates is still present even if mortality data from tropical African nations are excluded, and (2) reports that COVID-19 seroprevalence was high among Kenyan blood donors relative to COVID-19 case prevalence suggesting a high ratio of asymptomatic to symptomatic cases.^31^ Finally, we did not adjust our analyses based on national differences in date of earliest confirmed COVID-19 cases, however we do not believe that this factor would significantly influence our results given the extended duration of the pandemic period studied.

## Supporting information

in accordance with the PRISMA guideline

(See Supplementary Methods and Supplementary Table 1)

## Data Availability

All data are available and will be shared by request from any qualified investigator.

## Conclusions

Our meta-analysis points to an unprecedented conclusion: headache arising secondary to an infection is not a “non-specific” symptom, but rather it may be a marker of enhanced survival. We find that patients reporting headache in the setting of COVID-19 are at reduced risk of death, but that nations with high prevalence of headache disorders are at higher risk of COVID-19 mortality and possibly a lower risk of influenza mortality. Further studies are needed to clarify the roles of headache disorders in the context of viral infections or post-viral syndromes (e.g. “Long-Haul COVID”). Defining specific headache mechanisms that could enhance viral survival represents not only an opportunity for the discovery of improved COVID-19 therapeutics, but also for understanding whether, and how, primary headache disorders may be adaptive.

## Standard Protocol Approvals, Registrations, and Patient Consents

The study was conducted on available international datasets and on studies published in literature. No individual patients were included in the study.

## Data Availability

All data are available and will be shared by request from any qualified investigator.

## Author contributions

Each author (1) made substantial contributions to the conception and design of the work; acquisition, analysis, and interpretation of data; and drafted and substantively revised it, (2) approved the submitted version; and (3) agrees to be personally accountable for the author’s own contributions and to ensure that questions related to the accuracy or integrity of any part of the work, even ones in which the author was not personally involved, are appropriately investigated, resolved, and the resolution documented in the literature.

## Acknowledgements

We acknowledge the collaboration to this work of Dr. Rubio-Rivas (Department of Internal Medicine, Bellvitge University Hospital, Bellvitge Biomedical Research Institute-IDIBELL, University of Barcelona, Barcelona, Spain). We are also grateful to the hundreds of other investigators that compiled the multiple datasets that we relied upon in our analyses. We also especially remember all who have died due to COVID-19, and those who suffered or are still suffering from it. We acknowledge the efforts of all people, especially healthcare professionals, in the fight against COVID-19.

