## Supplementary material for "The impact of headache disorders on COVID-19 survival: a world population-based analysis": (See Supplementary Methods and Supplementary Table 1)

**Table of contents**

**Supplementary Methods.**

**Supplementary Table 1.** Search strategy performed on PubMed database search

**Supplementary Table 2.** Variables included in the generalized additive model

**Supplementary Table 3.** Studies selected for the meta-analysis

**Supplementary Table 4.** Pooled prevalence of symptoms and signs among COVID-19 inpatients

**Supplementary Table 5.** Risk of other symptoms in COVID-19 patients in studies where headache symptom is reported

**Supplementary Table 6.** Risk of other comorbid diseases in COVID-19 Patients in studies where headache symptom is reported

**Supplementary Table 7.** Sensitivity analysis of headache RR in COVID-19 patients

**Supplementary Table 8.** Variables associated with COVID-19 mortality across 171 nations in the generalized additive single-factor model

**Supplementary Table 9.** Variables associated with COVID-19 mortality across 171 nations in the generalized additive multi-factor model

**Supplementary Table 10.** Variables associated to COVID-19 mortality across 171 nations in the generalized additive multi-factor model in two different cohorts: Tropical (n=78) and non-tropical (n=93) nations.

**Supplementary Table 11.** Variables associated to Influenza mortality (2017) across 171 nations in the generalized additive multi-factor model

**Supplementary Figure 1.** The PRISMA flow diagram of the meta-analysis.

**Supplementary Figure 2.** Funnel plot on the risk of headache in COVID-19 patients after excluding outliers and over-influencer publications (n=41/48).

**Supplementary Figure 3.** Non-linear relationship (Spearman’s rho) between study variables.

**Supplementary Figure 4.** Influenza mortality (per million population) 2017 distribution across 171 nations based on diseases prevalence (according to GBD2019): (A) Urban population rate; (B) Population’s median age; (C) Population’s female rate; (D) National wealth; (E) Government effectiveness; (F) Capital’s latitude; (G) Headache disorders; (H) Diabetes mellitus; (I) Chronic kidney diseases; (J) Cirrhosis and Chronic liver diseases; (K) Chronic respiratory diseases and (L) Cardiovascular diseases.

**Meta-analysis references**

**Supplementary Methods**

(1) **Meta-analysis**

*Search strategy*

The search terms were “COVID-19” (OR “COVID-19 disease” OR “SARS-CoV-2” OR “SARS-CoV2” OR “nCoV”), “mortality” (OR “deceased” OR “died” OR “death” OR “fatality rate” OR “risk factor” OR “fatal” OR “dying”), “hospitalization” (OR “hospitalized” OR “hospitalized” OR “admission” OR “in-hospital”), “recovered” (OR “survived” OR “surviving” OR “survivors” OR “prognosis” OR “prognostic”) and “headache” (Supplementary Table 1). There was no restriction on study design, language nor laboratory confirmation of COVID-19 diagnosis. Studies were included if they clearly presented in their results or in the supplementary material: (1) study design; (2) COVID-19 confirmation method; (3) patient’s demographics; (4) ratio of COVID-19 survivors and non-survivors and (5) the presence of headache symptom in both cohorts.

*Study selection and data extraction*

Three investigators (VG, EC and PP-R) examined all titles and abstracts, and obtained full text of potentially relevant papers and data extraction was done by VG. For all eligible studies, we extracted information on study country, study size, COVID-19 confirmation, patients’ characteristics, including demographics and percentage of individuals with presence of other COVID-19 accompanying symptoms (anosmia, cough, diarrhea, dyspnea, fever, myalgia and nausea or vomiting), comorbidities (cardiovascular diseases, chronic kidney diseases, chronic liver diseases, chronic respiratory and diabetes) and our COVID-19 confirmation outcome of interest, the presence of headache.

*Data analysis*

In case of higher heterogeneity (I2 > 75%) in the RR analysis, if the publication’s CI did not overlap with the CI of the pooled effect, we considered these studies as outliers. Influence analyses of effect size between publications were also computed in order to assess whether the influence of a particular publication distorted the overall pooled effect. Other strategies considered in the sensitivity analysis were excluding small studies (n < 250); excluding studies lacking validated COVID-19 confirmation methods, and considering only prospective studies. Finally, publication bias was assessed through visual inspection (funnel plot) and significance test (Egger’s test). All of the statistical analysis and plots were generated using metaprop (version 2.4-0), meta (version 4.15-1) and dmetar (version 0.0.9) packages of R (version 4.0.3) software.

(2) **COVID-19 mortality-related factors: headache disorders**

Generalized additive models (GAMs) are regression models that can make more reasonable nonlinear fittings than traditional statistical models (Generalized Linear Models, GLMs). In particular, (i) they allow explanatory variables to be added into the model by function; and (ii) they can directly deal with the complex nonlinear relationship between multiple response and explanatory variables. GAMs require fewer data and can be applied to a variety of distribution types, like the Poisson distribution in this study. In the single-factor model, we evaluated all explanatory variables individually and we measured their significance (P value), deviance explained (%), adjusted R2 and the Akaike Information Criterion (AIC) for each variable. We used the adjusted R2 and variance interpretation rate (deviance explained) to evaluate the quality of the fitted GAM. We used the mgcv and car package in R 4.0.3 to analyze the GAM.

**Supplementary Table 1. Search strategy performed on PubMed database search.**

**Search Terms**

((((((((((((COVID-19[Title]) OR COVID19[Title]) OR coronavirus[Title]) OR nCoV[Title]) OR SARS-CoV-2[Title]) OR SARS-CoV2[Title])) AND

(((((((((((((((((((((mortality[Title/Abstract]) OR deceased [Title/Abstract]) OR died [Title/Abstract]) OR recovered [Title/Abstract]) OR death [Title/Abstract]) OR fatility rate [Title/Abstract]) OR risk factor [Title/Abstract]) OR fatal [Title/Abstract]) OR hospitalization [Title/Abstract]) OR survived [Title/Abstract]) OR surviving [Title/Abstract]) OR hospitalized [Title/Abstract]) OR non-survivors [Title/Abstract]) OR admission [Title/Abstract]) OR in-hospital [Title/Abstract]) OR hospitalised [Title/Abstract]) OR dying [Title/Abstract]) OR prognosis [Title/Abstract]) OR c [Title/Abstract]) OR headache [Title/Abstract])) AND ("2020/04/01"[Publication Date] : "3000"[Publication Date]))

**Supplementary Table 2. Variables included in the generalized additive model.**

| **Variable Name** | **Variable Description** | **Source Data** |
| --- | --- | --- |
| Population Density | People per km^2^ of land area | Our World in Data^1^ |
| Urban Population Rate | Percentage of population urbanized | CIA^2^ |
| Median age | Country’s median age (years) |  |
| Female rate | Percentage of female population | Worldwide Governance Indicators^3^ |
| Wealth | GDP per capita (US dollars) | Worldometer^4^ |
| Government effectiveness | World Bank Government Effectiveness Index (2019) | Worldwide Governance Indicators^3^ |
| Pandemic preparedness | Global Health Security Index (2020) | GHS Index^5^ |
| Pandemic response | Government Response Stringency Index (Max. valued obtained in 2020) | Our World in Data^1^ |
| Latitude of country’s capital | Degrees | Wikipedia^6^ |
| Headache disorders prevalence | Prevalence/100k (2019), age standardized | Global Health Data Exchange^7^ |
| Migraine prevalence | Prevalence/100k (2019), age standardized |  |
| Tension-type headache prevalence | Prevalence/100k (2019), age standardized |  |
| Diabetes prevalence | Prevalence/100k (2019), age standardized |  |
| Chronic kidney diseases prevalence | Prevalence/100k (2019), age standardized |  |
| Chronic liver diseases prevalence | Prevalence/100k (2019), age standardized |  |
| Chronic respiratory diseases prevalence | Prevalence/100k (2019), age standardized |  |
| Cardiovascular diseases prevalence | Prevalence/100k (2019), age standardized |  |
| Cumulative COVID-19 deaths | Prevalence/1m (14^th^ December 2020) | Our World in Data^1^ |
| 2017 Influenza death rates | Prevalence/100k (2017) | (36) |

^1^https://ourworldindata.org

^2^https://www.cia.gov

^3^https://info.worldbank.org

^4^https://www.worldometers.info

^5^https://www.ghsindex.org

^6^https://en.wikipedia.org/wiki/List_of_national_capitals_by_latitude

^7^http://ghdx.healthdata.org

**Supplementary Table 3. Studies selected for the meta-analysis.**

| **Author** | **Research**  **Timing** | **Design** | **Country** | **Region** | **All COVID19 Confirmation** | **Sample Size** | **Gender Predominance** | **Median Age** | **REF** |
| --- | --- | --- | --- | --- | --- | --- | --- | --- | --- |
| Aksel G et al. | Prospective | Cohort Study | Turkey | Europe | Yes | 168 | Male | <60 years | (1) |
| Bellan M et al. | Retrospective | Cohort Study | Italy | Europe | Yes | 407 | Male | ≥70 years | (2) |
| Berenguer J et al. | Retrospective | Cohort Study | Spain | Europe | Yes | 4,035 | Male | 60-69 years | (3) |
| Caronna E et al. | Prospective | Cohort Study | Spain | Europe | No | 130 | Female | <60 years | (4) |
| Chen F et al. | Prospective | Cohort Study | China | Asia | Yes | 660 | Female | <60 years | (5) |
| Chen L et al. | Retrospective | Cohort Study | China | Asia | Yes | 1,859 | Male | <60 years | (6) |
| Chen R et al. | Retrospective | Cohort Study | China | Asia | Yes | 1,590 | Male | <60 years | (7) |
| Chen T et al. | Retrospective | Cohort Study | China | Asia | Yes | 274 | Male | 60-69 years | (8) |
| Cheng A et al. | Retrospective | Cohort Study | China | Asia | Yes | 305 | Male | 60-69 years | (9) |
| Deng Y et al. | Retrospective | Cohort Study | China | Asia | Yes | 225 | Male | <60 years | (10) |
| De Souza C et al. | Retrospective | Cross-Sectional | Brazil | South America | No | 9,807 | Female | 60-69 years | (11) |
| Du RH et al. | Retrospective | Cohort Study | China | Asia | No | 179 | Male | <60 years | (12) |
| Emara DM et al. | Retrospective | Cohort Study | Egypt | Africa | Yes | 120 | Male | <60 years | (13) |
| Gao S et al. | Retrospective | Cohort Study | China | Asia | Yes | 210 | Female | ≥70 years | (14) |
| Garibaldi BT et al. | Prospective | Cohort Study | US | North America | Yes | 832 | Male | 60-69 years | (15) |
| Gil-Rodrigo A et al. | Prospective | Cohort Study | Spain | Europe | Yes | 1,000 | Male | 60-69 years | (16) |
| Homayounieh F et al. | Retrospective | Cohort Study | Iran | Africa | Yes | 90 | Male | <60 years | (17) |
| Khalil K et al. | Prospective | Cohort Study | UK | Europe | Yes | 220 | Male | 60-69 years | (18) |
| Li J et al. | Retrospective | Cohort Study | China | Asia | Yes | 161 | Female | <60 years | (19) |
| Li M et al. | Retrospective | Cohort Study | China | Asia | Yes | 245 | Female | <60 years | (20) |
| Li X et al. | Retrospective | Cohort Study | US | North America | Yes | 1,022 | Male | 60-69 years | (21) |
| Ma X et al. | Retrospective | Cohort Study | China | Asia | Yes | 523 | Male | <60 years | (22) |
| Marengoni A et al. | Retrospective | Cohort Study | Italy | Europe | Yes | 165 | Male | 60-69 years | (23) |
| Martin-Moro F et al. | Retrospective | Cohort Study | Spain | Europe | No | 34 | Male | ≥70 years | (24) |
| Mendes A et al. | Retrospective | Cohort Study | Switzerland | Europe | No | 235 | Female | ≥70 years | (25) |
| Moon SS et al. | Retrospective | Cohort Study | Korea | Asia | Yes | 348 | Female | <60 years | (26) |
| Park JG et al. | Retrospective | Cohort Study | Korea | Asia | Yes | 289 | Female | ≥70 years | (27) |
| Rana MS et al. | Retrospective | Cohort Study | Pakistan | Africa | Yes | 100 | Male | <60 years | (28) |
| Rivera-Izquierdo M et al. | Retrospective | Cohort Study | Spain | Europe | Yes | 131 | Male | 60-69 years | (29) |
| Rodriguez-Molinero A et al. | Prospective | Cohort Study | Spain | Europe | Yes | 418 | Male | 60-69 years | (30) |
| Rodriguez-Nava G et al. | Retrospective | Cohort Study | US | North America | Yes | 313 | Male | 60-69 years | (31) |
| Rubio-Rivas M et al. | Retrospective | Cohort Study | Spain | Europe | Yes | 12,066 | Male | 60-69 years | (32) |
| Soares RDCM et al. | Retrospective | Cohort Study | Brazil | South America | No | 1,152 | Male | 60-69 years | (33) |
| Tomlins J et al. | Retrospective | Cohort Study | UK | Europe | Yes | 95 | Male | ≥70 years | (34) |
| Trigo J et al. | Retrospective | Cohort Study | Spain | Europe | Yes | 576 | Male | 60-69 years | (35) |
| Van Halem K et al. | Retrospective | Cohort Study | Belgium | Europe | Yes | 319 | Male | ≥70 years | (37) |
| Varol Y et al. | Retrospective | Cohort Study | Turkey | Europe | Yes | 383 | Male | <60 years | (38) |
| Vena A et al. | Retrospective | Cohort Study | Italy | Europe | Yes | 317 | Male | ≥70 years | (39) |
| Wang D et al. | Retrospective | Cohort Study | China | Asia | Yes | 107 | Male | <60 years | (40) |
| Wang L et al. | Retrospective | Cohort Study | China | Asia | Yes | 339 | Female | 60-69 years | (41) |
| Wang ZH et al. | Retrospective | Cohort Study | China | Asia | Yes | 59 | Male | 60-69 years | (42) |
| Yan Y et al. | Retrospective | Cohort Study | China | Asia | Yes | 193 | Male | 60-69 years | (43) |
| Yang X et al. | Retrospective | Cohort Study | China | Asia | Yes | 52 | Male | <60 years | (44) |
| Yu Z et al. | Retrospective | Cohort Study | China | Asia | Yes | 141 | Female | ≥70 years | (45) |
| Zhang J et al. | Retrospective | Cohort Study | China | Asia | Yes | 663 | Female | <60 years | (46) |
| Zhang L et al. | Retrospective | Cohort Study | China | Asia | Yes | 319 | Female | <60 years | (47) |
| Zhao Z et al. | Retrospective | Cohort Study | US | North America | Yes | 480 | Male | 60-69 years | (48) |
| Zhu J et al. | Retrospective | Cohort Study | US | North America | Yes | 181 | Male | <60 years | (49) |

**Supplementary Table 4. Pooled prevalence of symptoms and signs among COVID-19 inpatients**

| Symptoms | Symptom prevalence  [95% CI] | Number of studies included^*^ | Total number of COVID-19 patients | Between-study heterogeneity | | Publication bias, Egger’s test  (*P* value) |
| --- | --- | --- | --- | --- | --- | --- |
|  |  |  |  | *I^2^* | *P* value |  |
| Anosmia | 0.066 [0.041-0.104] | 15/16 | 9,128 | 97% | **<0.001** | 0.276 |
| Cough | 0.650 [0.612-0.687] | 22/43 | 11,309 | 93% | **<0.001** | 0.174 |
| Diarrhoea | 0.113 [0.084-0.149] | 23/35 | 22,121 | 97% | **<0.001** | 0.802 |
| Dyspnoea | 0.419 [0.339-0.503] | 22/40 | 6,506 | 98% | **<0.001** | 0.112 |
| Fever (>37.3 ºC) | 0.779 [0.720-0.829] | 34/42 | 27,068 | 99% | **<0.001** | 0.136 |
| Headache | 0.097 [0.078-0.120] | 41/48 | 30,236 | 97% | **<0.001** | 0.275 |
| Myalgia | 0.176 [0.146-0.211] | 34/38 | 25,581 | 97% | **<0.001** | 0.446 |
| Nausea or Vomiting | 0.103 [0.077-0.137] | 27/30 | 14,191 | 97% | **<0.001** | 0.109 |

^*^Studies with extreme effect sizes were discarded in order to obtain an unbiased publication effect (Egger’s test).

In **bold** are marked *P values* < 0.05

**Supplementary Table 5. Risk of symptoms among COVID-19 inpatients, in studies where headache symptom is reported**

| COVID-19 symptom | Test for overall effect  (random-model) | Comorbidity-COVID19 RR  [95% CI] | Number of studies included^*^ | Between-study heterogeneity | | Publication bias, Egger’s test (P value) |
| --- | --- | --- | --- | --- | --- | --- |
|  |  |  |  | I^2^ | P value |  |
| Anosmia | t = 8.76  **p<0.001** | 3.183 [2.397-4.226] | 15/16 | 0% | 0.890 | 0.560 |
| Cough | t = 1.71  p = 0.102 | 1.036 [0.992-1.081] | 22/43 | 24% | 0.147 | 0.148 |
| Diarrhea | t = 1.09  p = 0.287 | 1.078 [0.935-1.242] | 23/35 | 18% | 0.214 | 0.650 |
| Dyspnea | t = -12.61  **p<0.001** | 0.662 [0.618-0.708] | 22/40 | 25% | 0.142 | 0.550 |
| Fever (>37.3 º0C) | t = 1.65  p = 0.109 | 1.025 [0.994-1.058] | 34/42 | 46% | **0.002^†^** | 0.352 |
| Headache | t = 10.75  **p<0.001** | 2.178 [1.882-2.520] | 41/48 | 16% | 0.175 | 0.733 |
| Myalgia | t = 7.75  **p<0.001** | 1.574 [1.398-1.773] | 34/38 | 17% | 0.200 | 0.859 |
| Nausea or Vomiting | t = 4.87  **p<0.001** | 1.558 [1.292-1.879] | 27/30 | 22% | 0.158 | 0.604 |

^*^Studies with extreme effect sizes (outliers) and studies with higher influence on overall effect (Leave-One-Out influence analysis) were discarded in order to obtain a homogenized between-studies effect

**^†^**All models exploring fever as a RR resulted statistically significant heterogeneity between studies (there were studies where fever was reported at patient’s admission, other studies reported fever as a history symptom and the rest of studies did not specify the moment in which fever was considered)

In **bold** are marked *P values* < 0.05

**Supplementary Table 6. Risk of comorbid diseases in COVID-19 inpatients, in studies where headache symptom is reported**

| Comorbidity | Test for overall effect  (random-model) | Comorbidity-COVID19 RR  [95% CI] | Number of studies included^*^ | Between-study heterogeneity | | Publication bias, Egger’s test (P value) |
| --- | --- | --- | --- | --- | --- | --- |
|  |  |  |  | I^2^ | P value |  |
| Cardiovascular diseases | t = -13.41  **p<0.001** | 0.405 [0.352-0.466] | 23/34 | 30% | 0.090 | 0.179 |
| Chronic kidney diseases | t = -11.78  **p<0.001** | 0.338 [0.280-0.408] | 31/34 | 34% | 0.090 | 0.985 |
| Chronic liver diseases | t = -5.72  **p<0.001** | 0.530 [0.419-0.670] | 18/18 | 0% | 0.603 | 0.629 |
| Chronic respiratory diseases | t = -10.13  **p<0.001** | 0.519 [0.455-0.592] | 33/38 | 15% | 0.223 | 0.645 |
| Diabetes | t = -9.75  **p<0.001** | 0.613 [0.553-0.679] | 31/38 | 28% | 0.080 | 0.969 |

^*^Studies with extreme effect sizes (outliers) and studies with higher influence on overall effect (Leave-One-Out influence analysis) were discarded in order to obtain a homogenized between-studies effect

In **bold** are marked *P values* < 0.05

**Supplementary Table 7. Sensitivity analysis of headache relative risk (RR) among COVID-19 inpatients**

| Strategy | Test for overall effect  (random-model) | Headache-COVID19 RR  [95% CI] | Between-study heterogeneity | | Number of studies included | Publication bias, Egger’s test (P value) |
| --- | --- | --- | --- | --- | --- | --- |
|  |  |  | I^2^ | P value |  |  |
| Including all studies | t = 5.18  **p<0.001** | 1.876 [1.469-2.390] | 79% | **<0.001** | 48/48 | 0.787 |
| Excluding outliers and over-influencer studies | t = 10.75  **p<0.001** | 2.178 [1.882-2.520] | 16% | 0.175 | 41/48 | 0.733 |
| Excluding retrospective studies | t = 3.55  **p=0.012** | 3.380 [1.459-7.828] | 53% | **0.050** | 7/48 | 0.280 |
| Excluding studies without COVID-19 confirmation method | t = 5.18  **p<0.001** | 1.917 [1.489-2.469] | 76% | **<0.001** | 44/48 | 0.457 |
| Excluding small studies (n < 250 patients) | t = 7.26  **p<0.001** | 2.251 [1.790-2.829] | 65% | **<0.001** | 26/48 | 0.974 |

In **bold**, *P values* < 0.05

**Supplementary Table 8. Variables associated with COVID-19 mortality across 171 nations in the generalized additive single-factor model**

| Smoothed variable | Deviance explained (%) | Edf | Ref.df | *P* value | *F* value | Adjust R^2^ | AIC |
| --- | --- | --- | --- | --- | --- | --- | --- |
| **Headache disorders prevalence** | 37.8 | 8.675 | 8.969 | <2e-16^**^ | 10.68 | 0.345 | 2421.46 |
| **Latitude of country’s capital** | 34.8 | 7.113 | 8.173 | <2e-16^**^ | 9.99 | 0.319 | 2426.49 |
| **Migraine prevalence** | 32.0 | 8.262 | 8.843 | <2e-16^**^ | 8.38 | 0.286 | 2435.79 |
| **Median age** | 31.1 | 4.811 | 5.920 | <2e-16^**^ | 12.12 | 0.291 | 2431.25 |
| **Chronic liver diseases prevalence** | 28.6 | 4.627 | 5.707 | <2e-16^**^ | 11.12 | 0.266 | 2437.02 |
| **Tension-type headache prevalence** | 26.7 | 4.471 | 5.520 | <2e-16^**^ | 10.41 | 0.247 | 2441.18 |
| **Wealth** | 24.7 | 6.922 | 7.903 | 9.10e-07^**^ | 6.40 | 0.215 | 2450.62 |
| **Pandemic preparedness** | 19.8 | 1.092 | 1.179 | <2e-16^**^ | 34.38 | 0.193 | 2449.83 |
| **Urban population rate** | 17.3 | 1.000 | 1.000 | <2e-16^**^ | 35.33 | 0.168 | 2454.88 |
| **Government effectiveness** | 16.5 | 2.915 | 3.663 | 1.01e-05^**^ | 8.45 | 0.150 | 2460.37 |
| **Cardiovascular diseases prevalence** | 14.6 | 5.153 | 6.285 | 8.13e-4^**^ | 4.00 | 0.119 | 2468.70 |
| **Chronic respiratory diseases prevalence** | 13.6 | 2.948 | 3.700 | 1.17e-4^**^ | 6.86 | 0.121 | 2466.15 |
| **Pandemic response** | 7.8 | 1.117 | 1.225 | 0.001^**^ | 12.04 | 0.071 | 2473.74 |
| Chronic kidney diseases prevalence | 7.4 | 5.521 | 6.663 | 0.164 | 1.57 | 0.043 | 2483.20 |
| **Diabetes prevalence** | 6.9 | 2.723 | 3.402 | 0.026^*^ | 3.06 | 0.054 | 2478.48 |
| Female rate | 2.4 | 1.604 | 2.008 | 0.183 | 1.73 | 0.015 | 2484.32 |
| Population density | 0.4 | 1.000 | 1.000 | 0.388 | 0.75 | 0.000 | 2486.58 |

Estimated degree of freedom (Edf), degree of reference (Ref. df), *P* value, *F* value (measurement of the relative importance of smoothed variable), deviance explained (%), adjusted R^2^, and Akaike information criterion (AIC) for the smoothed variables (including population density, urban population rate, median age, female rate, wealth, government effectiveness, pandemic preparedness, pandemic response, latitude of country’s capital and comorbidities prevalence) in the single-factor model for COVID-19 mortality.

In **bold** are statistically significant smoothed variables in the single-factor model.

^*^This indicates *P* values < 0.05

^**^This indicates *P* values < 0.01

**Supplementary Table 9. Variables associated with COVID-19 mortality across 171 nations in the generalized additive multi-factor model**

| Smoothed variable | Edf | Ref.df | *P* value | *F* value | Deviance explained (%) | Adjust R^2^ | AIC |
| --- | --- | --- | --- | --- | --- | --- | --- |
| **Urban population rate** | 1.854 | 2.346 | 2.61e-4^**^ | 8.14 | 65.5 | 0.591 | 2356.85 |
| Median age | 1.000 | 1.000 | 0.346 | 0.90 |  |  |  |
| **Wealth** | 1.000 | 1.000 | 4.83e-4^**^ | 12.8 |  |  |  |
| Pandemic preparedness | 1.980 | 2.482 | 0.204 | 1.53 |  |  |  |
| Pandemic response | 1.000 | 1.000 | 0.217 | 1.54 |  |  |  |
| **Latitude of country’s capital** | 8.501 | 8.918 | <2e-16^**^ | 6.84 |  |  |  |
| **Migraine prevalence** | 8.262 | 8.843 | 0.009^*^ | 3.84 |  |  |  |
| Diabetes prevalence | 1.000 | 1.000 | 0.216 | 1.54 |  |  |  |
| Chronic liver diseases prevalence | 1.000 | 1.000 | 0.161 | 1.98 |  |  |  |
| Chronic respiratory diseases prevalence | 4.147 | 5.139 | 0.201 | 1.55 |  |  |  |
| Cardiovascular diseases prevalence | 3.096 | 3.930 | 0.051 | 2.367 |  |  |  |

Estimated degree of freedom (Edf), degree of reference (Ref. df), *P value*, *F* value, deviance explained (%), adjusted R^2^, and Akaike information criterion (AIC)

for the smoothed variables in the multi-factor model for COVID-19 mortality.

In the multi-factor model, only statistically significant variables from the single-factor were included. Government effectiveness was not considered because it presented a statistically significant high correlation (r≥0.70; p<0.05) with national wealth, pandemic preparedness and pandemic response. Headache disorders prevalence, migraine prevalence and tension-type headache prevalence were also highly correlated: Hence, we modelled individually in the multi-factor model and we selected the best model based on AIC (i.e., best model considering migraine prevalence instead of headache disorders prevalence or tension-type headache prevalence).

In **bold** are statistically significant smoothed variables in the multi-factor model.

^*^This indicates *P values* < 0.05

^**^This indicates *P values* < 0.01

**Supplementary Table 10. Variables associated with COVID-19 mortality across 171 nations in the generalized additive multi-factor model in two different cohorts: Tropical (n=78) and non-tropical (n=93) nations.**

| Smoothed variable | Edf | Ref.df | P value | F value | Deviance explained (%) | Adjust R^2^ | AIC |
| --- | --- | --- | --- | --- | --- | --- | --- |
| Non-Tropical countries (n=93) | | | | | | | |
| **Urban population rate** | 1.000 | 1.000 | 0.009^**^ | 7.07 | 68.3 | 0.575 | 1305.77 |
| **Median age** | 4.131 | 5.114 | 0.023^*^ | 2.82 |  |  |  |
| Pandemic preparedness | 1.000 | 1.000 | 0.737 | 0.11 |  |  |  |
| **Latitude of country’s capital** | 7.738 | 8.552 | 2.39e-5^**^ | 5.22 |  |  |  |
| **Migraine prevalence** | 1.712 | 2.133 | 0.004^**^ | 5.56 |  |  |  |
| Chronic kidney diseases prevalence | 5.752 | 6.872 | 0.156 | 1.61 |  |  |  |
| Chronic liver diseases prevalence | 1.000 | 1.000 | 0.498 | 0.46 |  |  |  |
| Chronic respiratory diseases prevalence | 1.000 | 1.000 | 0.435 | 0.61 |  |  |  |
| Tropical countries (n=78) | | | | | | | |
| **Urban population rate** | 7.235 | 7.917 | 0.001^**^ | 4.58 | 96.6 | 0.914 | 913.50 |
| **Median age** | 2.227 | 2.744 | 9.31e-4^**^ | 7.50 |  |  |  |
| **National wealth** | 8.347 | 8.711 | 2.15e-5^**^ | 7.33 |  |  |  |
| **Pandemic preparedness** | 8.740 | 8.944 | 1.24e-4^**^ | 5.79 |  |  |  |
| **Migraine prevalence** | 3.912 | 4.667 | 0.013^*^ | 3.79 |  |  |  |
| **Chronic kidney diseases prevalence** | 6.573 | 7.561 | 0.002^**^ | 4.26 |  |  |  |
| **Cardiovascular diseases prevalence** | 8.728 | 8.944 | <2e-16^**^ | 14.52 |  |  |  |
| Chronic liver diseases prevalence | 1.000 | 1.000 | 0.239 | 1.44 |  |  |  |

Estimated degree of freedom (Edf), degree of reference (Ref. df), P-value, F-value (which measures the relative importance of smoothed variable), deviance explained (%), adjusted R^2^, deviance contribution (%), and Akaike information criterion (AIC) for the smoothed variables in the multi-factor model between non-tropical and tropical countries. In the multivariate model, only statistically significant variables from the single-factor (tropical and non-tropical countries) were included. Headache disorders prevalence, migraine prevalence and tension-type headache prevalence were also highly correlated: hence, we modelled individually in the multi-factor model and we selected the best model based on AIC (model considering migraine prevalence).

In **bold** are marked statistically significant smoothed variables in the single-factor model.

^*^This indicates *P values* < 0.05; ^**^This indicates *P values* < 0.01

**Supplementary Table 11. Variables associated with Influenza mortality (2017) across 171 nations in the generalized additive multi-factor model**

| Smoothed variable | Edf | Ref.df | P value | F value | Deviance explained (%) | Adjust R^2^ | AIC |
| --- | --- | --- | --- | --- | --- | --- | --- |
| Urban population rate | 6.941 | 7.999 | 0.175 | 1.46 | 84.3 | 0.802 | 1578.66 |
| Median age | 1.324 | 1.577 | 0.121 | 1.85 |  |  |  |
| Wealth | 1.000 | 1.000 | 0.603 | 0.27 |  |  |  |
| Government effectiveness | 1.000 | 1.000 | 0.638 | 0.22 |  |  |  |
| **Latitude of country’s capital** | 7.815 | 8.610 | 6.52e-6^**^ | 5.41 |  |  |  |
| **Headache disorders prevalence** | 1.977 | 2.505 | 0.016^*^ | 4.08 |  |  |  |
| **Diabetes prevalence** | 4.031 | 4.967 | 3.49e-4^**^ | 4.96 |  |  |  |
| **Chronic kidney diseases prevalence** | 1.000 | 1.000 | 1.25e-5^**^ | 20.65 |  |  |  |
| **Chronic liver diseases prevalence** | 2.613 | 3.338 | 4.33e-4^**^ | 6.15 |  |  |  |
| Chronic respiratory diseases prevalence | 1.847 | 2.311 | 0.277 | 1.17 |  |  |  |
| Cardiovascular diseases prevalence | 5.107 | 6.200 | 0.065 | 1.99 |  |  |  |

Estimated degree of freedom (Edf), degree of reference (Ref. df), P value, F value, deviance explained (%), adjusted R^2^, and Akaike information criterion (AIC) for the smoothed variables in the multi-factor model for Influence mortality. In the multi-factor model, only statistically significant variables from the single-factor were included. Headache disorders prevalence, migraine prevalence and tension-type headache prevalence were highly correlated (r≥0.70; p<0.05): Hence, we modelled individually in the multi-factor model and we selected the best model based on AIC (best model considering tension-type headache prevalence instead of headache disorders prevalence or migraine prevalence).

In **bold** are marked statistically significant smoothed variables in the multi-factor model.

^*^This indicates *P value* < 0.001

^**^This indicates *P value* < 0.0001

**Supplementary Figure 1. The PRISMA flow diagram of the meta-analysis**

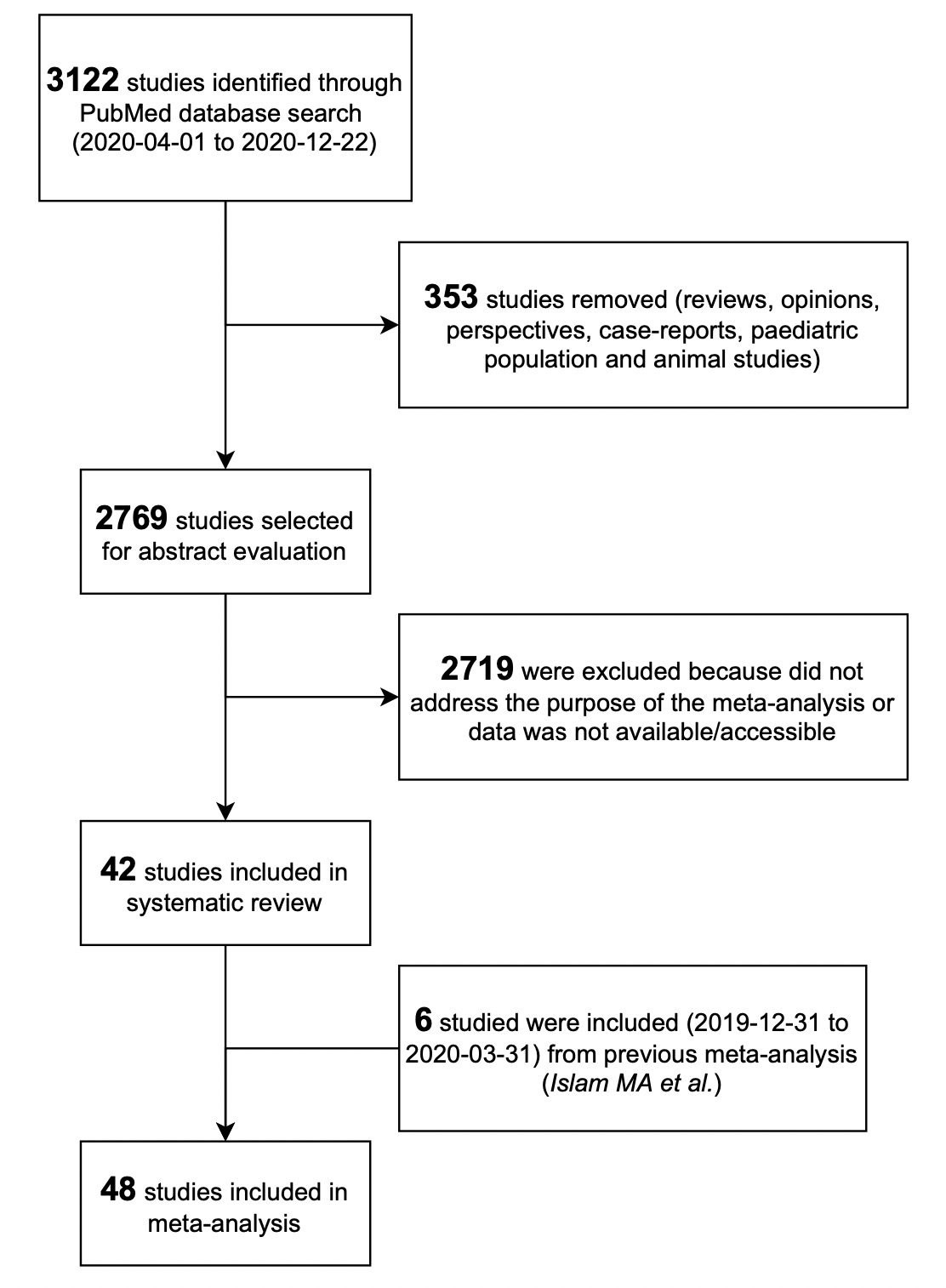

**Supplementary Figure 2. Funnel plot on the risk of headache among COVID-19 inpatients, after excluding outliers and over-influencer publications (n=41/48).**

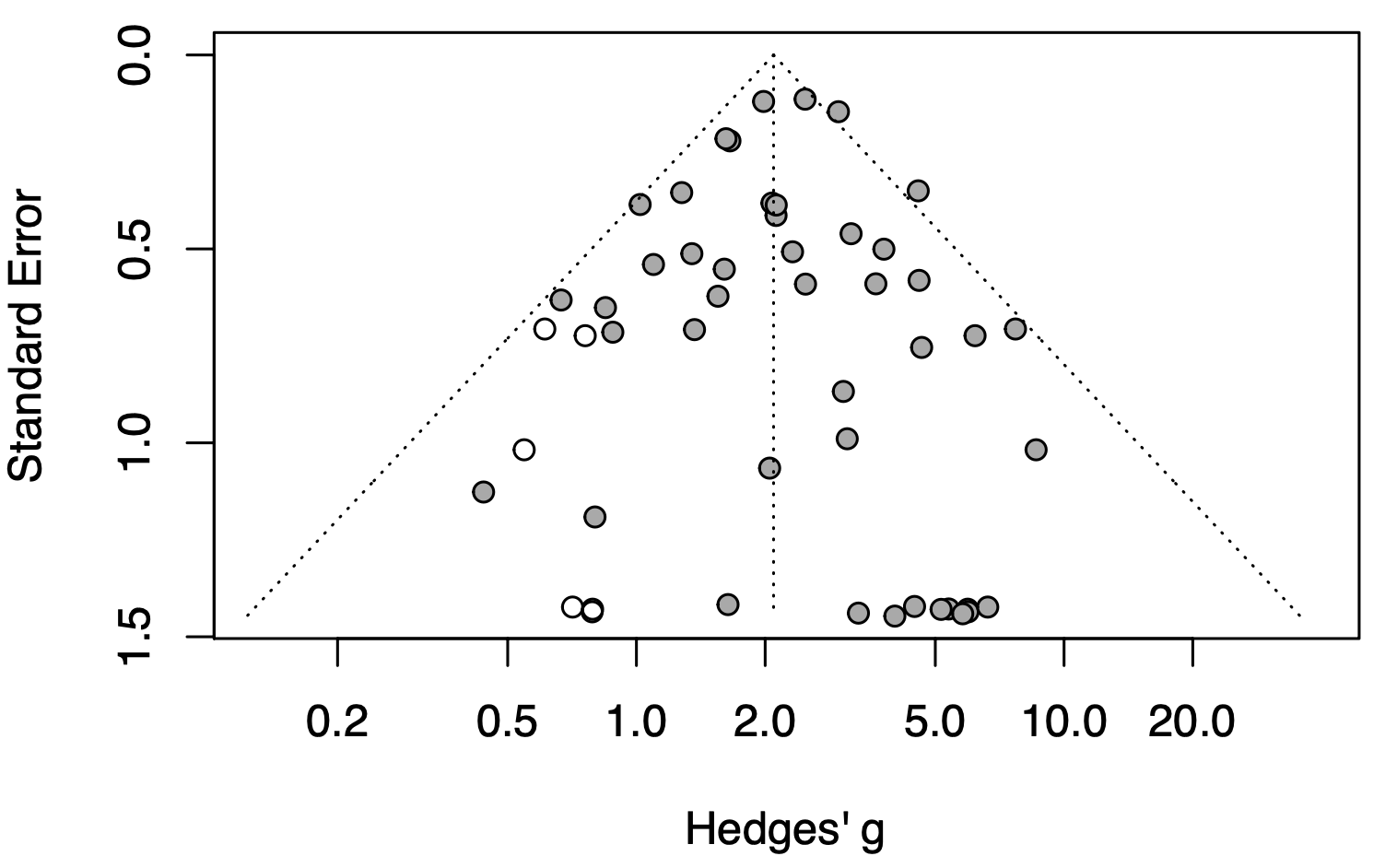

Egger’s test significance was *P value* = 0.733

**Supplementary Figure 3. Non-linear relationship (Spearman’s rho) between study variables.**

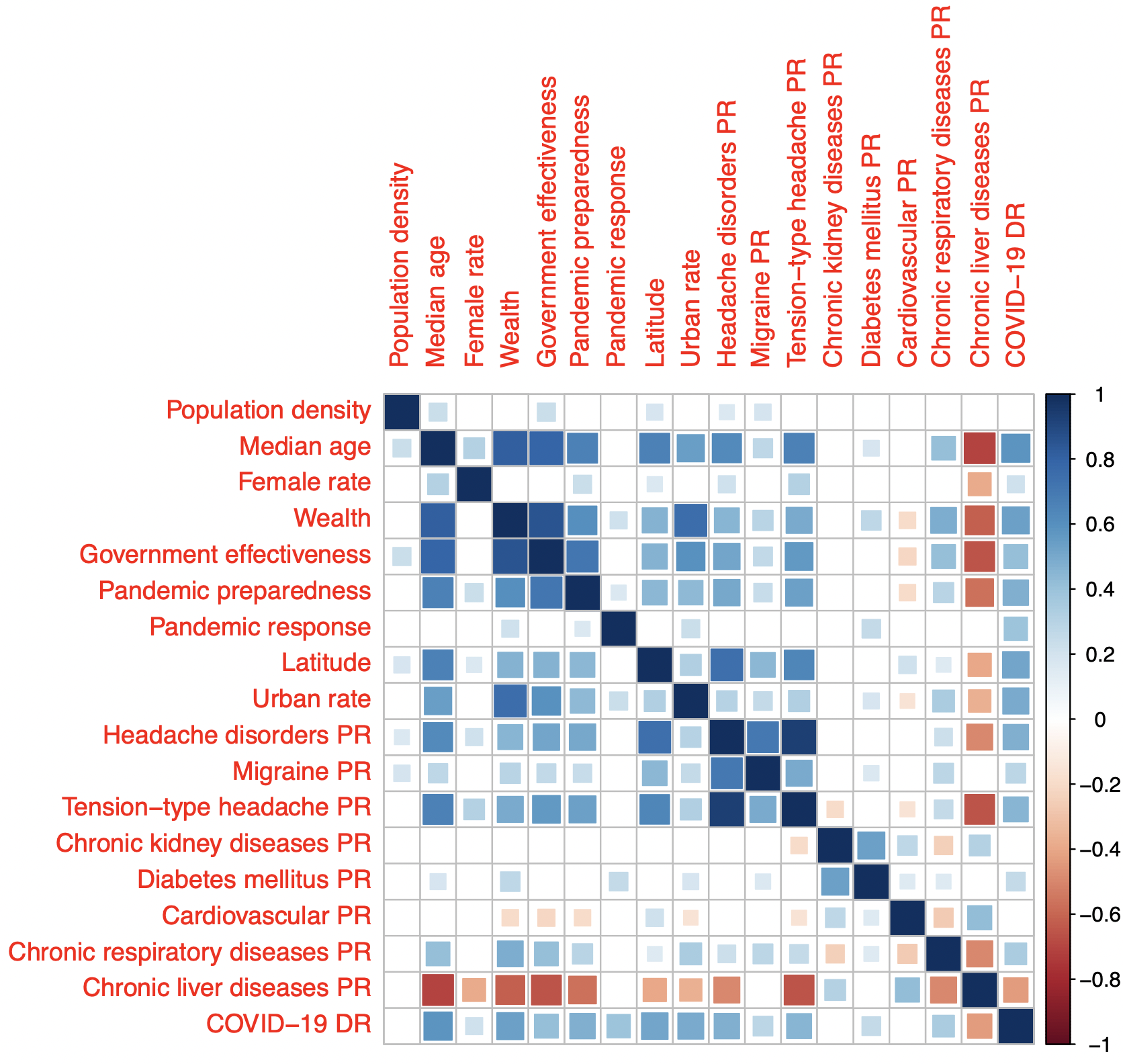

Blank cells mean non statistically significant correlation between each pair of variables. Blue indicates positive correlation and red indicates negative correlation between each pair of variables.

Abbreviations: PR: Prevalence; DR: Death Rate

**Supplementary Figure 4. Influenza mortality (deaths per 100K) 2017 distribution across 171 nations based on diseases prevalence (according to GBD2017): (A) Urban population rate; (B) Population’s median age; (C) Population’s female rate; (D) National wealth; (E) Government effectiveness; (F) Capital’s latitude; (G) Headache disorders; (H) Diabetes mellitus; (I) Chronic kidney diseases; (J) Cirrhosis and Chronic liver diseases; (K) Chronic respiratory diseases and (L) Cardiovascular diseases.**

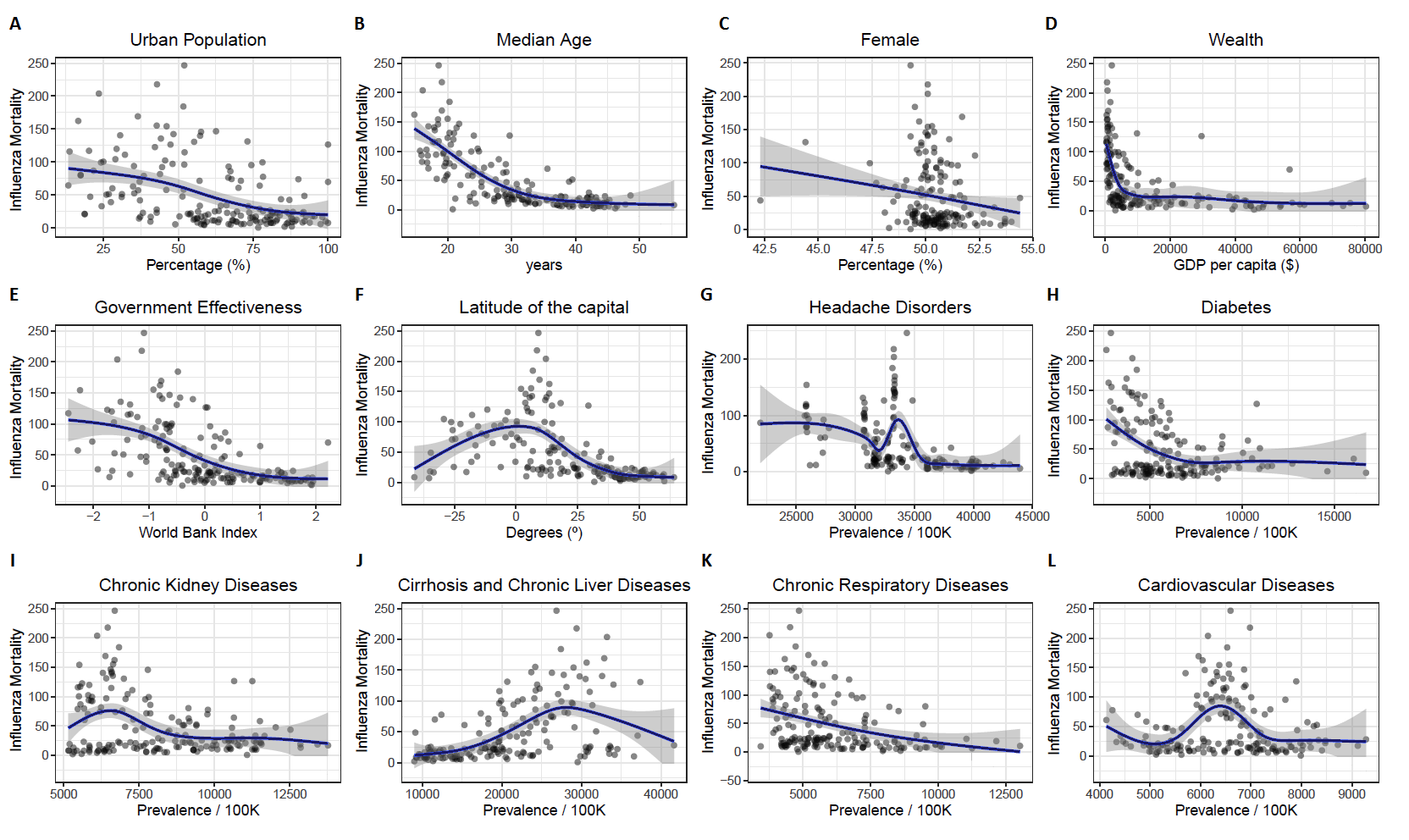

Abbreviations: GBD: Global Burden of Diseases

**Meta-Analysis references**
